# Behavioural changes in frontotemporal dementia and their cognitive and neuroanatomical correlates

**DOI:** 10.1101/2024.09.24.24314224

**Authors:** Matthew A. Rouse, Masud Husain, Peter Garrard, Karalyn Patterson, James B. Rowe, Matthew A. Lambon Ralph

**Author notes:** Correspondence to: Dr Matthew A. Rouse, MRC Cognition and Brain Sciences Unit, University of Cambridge, CB2 7EF, Cambridge, UK.

## Abstract

Behavioural changes are a central feature of frontotemporal dementia (FTD); they occur in both behavioural-variant (bvFTD) and semantic dementia (SD)/semantic-variant primary progressive aphasia subtypes. In this study, we addressed two current clinical knowledge gaps; (i) are there qualitative or clear distinctions between behavioural profiles in bvFTD and SD, and (ii) what are the precise roles of the prefrontal cortex and anterior temporal lobes in supporting social behaviour? Resolving these conundrums is crucial for improving diagnostic accuracy and for the development of targeted interventions to treat challenging behaviours in FTD. Informant questionnaires to assess behavioural changes included the Cambridge Behavioural Inventory-Revised and two targeted measures of apathy and impulsivity. Participants completed a detailed neuropsychological battery to permit investigation of the relationship between cognitive status (including social-semantic knowledge, general semantic knowledge and executive function) with behaviour change in FTD. To explore changes in regional grey matter volume, a subset of patients had structural MRI. Diagnosis-based group comparisons were supplemented by a transdiagnostic approach which encompassed the spectrum of bvFTD, SD and “mixed” or intermediate cases. Such an approach is sensitive to the systematic graded variation in FTD and allows the neurobiological underpinnings of behaviour change to be explored across an FTD spectrum. We found a wide range of behavioural changes across FTD. Although *quantitatively* more severe on average in bvFTD, as expected, the item-level analyses found no evidence for *qualitative* differences in behavioural profiles or “behavioural double dissociations” between bvFTD and SD. Comparisons of self and informant ratings revealed strong discrepancies in the perspective of the caregiver versus patient. Logistic regression revealed that neuropsychological measures had better discriminative accuracy for bvFTD versus SD than caregiver-reported behavioural measures. A principal component analysis of all informant questionnaire domains extracted three components, interpreted as reflecting: (1) apathy, (2) challenging behaviours and (3) activities of daily living. More severe apathy in both FTD subtypes was associated with (a) increased levels of impaired executive function and (b) anterior cingulate cortex atrophy. Questionnaire ratings of impaired behaviour did not correlate with either anterior temporal lobe atrophy or degraded social-semantic knowledge. Together, these findings highlight the presence of a wide range of behavioural changes in both bvFTD and SD, which vary by degree rather than quality. We recommend a transdiagnostic approach for future studies of the neuropsychological and neuroanatomical underpinnings of behavioural deficits in FTD.

## Introduction

Behavioural changes are a core manifestation of frontotemporal dementia (FTD) and have a considerable impact on both patients and their caregivers.^1–3^ They are classically associated with behavioural-variant FTD (bvFTD) and the prefrontal cortical atrophy in this condition^4–7^ although have also been linked to other brain areas and changes in connectivity.^8^ Behavioural changes are now recognised as common in semantic dementia (SD; encompassing semantic-variant primary progressive aphasia and ‘right’ semantic dementia or right temporal variant FTD)^9–13^ where pathology is centred on the anterior temporal lobes (ATLs) leading to degraded semantic memory.^14,15^ This study aimed to resolve two key current gaps in clinical knowledge. First, are the behavioural changes in bvFTD and SD largely the same or are there qualitatively distinct behavioural profiles? Identifying discriminative behaviours would improve management and expectations in clinic and improve bvFTD versus SD diagnostic accuracy. Such accuracy is particularly relevant for disease-modifying clinical trial design, as the two disorders are typically associated with different neuropathologies.^16^ Second, what are the precise contributions of the prefrontal cortex and the ATL in supporting social behaviour? Revealing the cognitive and neurobiological mechanisms underlying behaviour change in FTD is vital for informing the development of targeted pharmacological and behavioural interventions. To address these clinical conundrums, we explored the range of behavioural changes that are caused by FTD and the similarities and/or differences between FTD subtypes. Participants also completed extensive neuropsychological testing and structural MRI, to investigate the cognitive and neuroanatomical bases of changed behaviours. This calls for a transdiagnostic approach, including not only archetypal cases of bvFTD and SD but also “mixed” or intermediate cases that express prominent clinical features of both conditions, as part of a continuous clinical spectrum.^17^

People with FTD can behave in ways that reflect a loss of or disregard for social norms or etiquette, which can be misinterpreted as disinhibition or loss of empathy.^13,18^ Apathy and impulsivity are also common and co-occurring features of FTD and may exacerbate abnormal social behaviours.^19^ Behavioural disturbances are the hallmark of bvFTD and have been associated with structural, functional and neurochemical changes in the orbitofrontal cortex, inferior frontal gyrus, anterior cingulate cortex, insula and their connected subcortical structures.^20–24^ The frontopolar cortex is another area atrophied in bvFTD, with one theory proposing that this region supports representation of the long-term consequences of social behaviour.^25^

Behavioural changes are also common in SD, despite initial presentation with semantic and language deficits. Indeed, large cohort studies have reported similar rates of behaviour change in SD and bvFTD.^17,19,26,27^ Unlike bvFTD, the atrophy in SD is primarily centred on the ATLs.^15^ In their severest form, the co-occurring semantic and behavioural impairments are reminiscent of the Klüver-Bucy syndrome, which is characterised by a multimodal associative agnosia and chronic behavioural change following bilateral (but not unilateral) ATL ablation in macaques^28,29^ (and in rare human cases).^30^ Behavioural changes and prosopagnosia are commonly reported in SD patients with asymmetric rightward biased ATL atrophy^31–33^; this clinical observation has led to proposals that the right ATL has a specialised role for social-semantic knowledge^33,34^ and that these patients constitute a distinct clinical entity - the right temporal variant FTD.^31,33,35^ However, formal assessments have demonstrated that social deficits are also found in SD with left dominant atrophy.^31,36,37^ Recent investigations have found that degraded social-semantic knowledge is associated with *bilateral* ATL atrophy in FTD: there is no evidence for selective or greater social-semantic deficits in the presence of R>L atrophy or following selective right ATL resection for surgical treatment of epilepsy.^38,39^ Consequently, it appears that the left and right ATLs are both important for social behaviour and need to be investigated in FTD alongside prefrontal cortical regions.

We have proposed a neurocognitive model of impaired social behaviour in FTD - *controlled social-semantic cognition* (CS-SC).^8^ According to the CS-SC framework, impaired social behaviour in FTD results from damage to two distinct yet interactive components – (i) ATL-based *social-semantic knowledge*, and (ii) *social control*, which is supported by the prefrontal cortex. Social-semantic knowledge refers to our long-term database of the meaning of words, objects and behaviours acquired over our lifetimes and is critical to understand and generate appropriate behaviours across specific contexts.^8,34,40^ In a previous study, we demonstrated that social- and non-social-semantic deficits were highly correlated in FTD and were uniquely associated with *bilateral* ATL atrophy.^39^ Social control includes the selection, evaluation, decision-making and inhibition processes supported by the orbitofrontal and medial prefrontal cortices and interacts with ATL-based semantic representations to guide controlled social behaviour.^8^ A key hypothesis from the model is that the behavioural changes in SD result predominantly from a degradation of social-semantic knowledge following bilateral ATL atrophy, whereas the behavioural changes in bvFTD appear to result primarily from difficulties controlling social-semantic knowledge effectively to guide socially appropriate behaviour across changing contexts and scenarios.^8^

In this study, we measured the range of behavioural changes in FTD (spanning bvFTD, SD and intermediate cases) and their cognitive and neuroanatomical bases. We utilised the Cambridge Behavioural Inventory-Revised and two questionnaires of apathy and impulsivity, together with neuropsychological assessments and structural MRI. For the first time, we systematically explored the link between comprehension of a wide range of social concepts and behaviour change in the same FTD cohort. To achieve comprehensive coverage of behavioural changes in FTD, two additional informant questionnaires were applied alongside the CBI-R: the Cambridge Questionnaire for Apathy and Impulsivity Traits (CamQUAIT) and the Apathy-Motivation Index-Caregiver version (AMI-CG). The CamQUAIT measures apathy and impulsivity/challenging behaviours and was included because, unlike the CBI-R and other standard measures, the questionnaire was developed and validated specifically for use in the context of people with frontotemporal lobar degeneration. Apathy is a core feature of FTD, and considered to be multidimensional construct, so we included the AMI-CG as this questionnaire has been shown to be sensitive to subtypes of apathy, to explore (a) whether apathy subtypes could be disentangled in FTD, and (b) whether each subtype had separable neural substrates and neuropsychological correlates.

The inherently shared phenotypic variation in FTD and the highly correlated atrophy across frontotemporal areas means that inter-subgroup comparisons limit the ability to localise precise functions to specific brain regions, and they can also be blind to the patterns of phenotypic variation that occur across the FTD clinical spectrum (including intermediate FTD cases who do not fall neatly into one diagnostic category).^17,26,27,39,41^ Therefore, we also used a data-driven transdiagnostic approach which treats FTD as a spectrum where patients represent phenotypic points along a frontotemporal atrophy continuum,^42^ to supplement classical diagnosis-based comparisons with multivariate analytics.

## Materials and methods

### Participants

Forty-seven people with a clinical diagnosis of FTD were recruited from specialist clinics in the Cambridge Centre for Frontotemporal Dementia at the Cambridge University Hospitals NHS Trust (Addenbrookes) (*n* = 40), St George’s Hospital, London (*n* = 4) and John Radcliffe Hospital, Oxford (*n* = 3). Twenty-six people had a primary diagnosis of bvFTD^5^ whereas 21 met criteria for SD (encompassing both L>R and R>L patterns of temporal atrophy).^14,15^ For each participant, grey matter intensity values in the left and right ATL were extracted and linear regression models fitted using the control data, with each region of interest as the dependent variable and age, intracranial volume (ICV) and scanner site included as covariates. Each FTD patient’s data were entered into the model, and the residuals were used to calculate two indices: ATL magnitude (left + right) and ATL asymmetry (left – right). Of the SD group, 18 had L>R ATL atrophy and 3 had R>L ATL atrophy, an uneven balance that aligns with clinical experience. The distribution of graded ATL magnitude and asymmetry values for each FTD case are displayed in Supplementary Fig. 1.

Eighteen age-matched healthy participants were recruited from the MRC Cognition and Brain Sciences Unit, University of Cambridge. Most participants provided written informed consent obtained according to the Declaration of Helsinki. Where participants lacked capacity to consent, their next of kin was consulted using the ‘personal consultee’ process as established by UK law. Demographic and disease information is reported in Table 1. There was a significant difference in sex distribution found between groups, however no significant interactions between group and sex were detected for any group comparisons, indicating that sex had no influence on the findings.

**Table 1.**
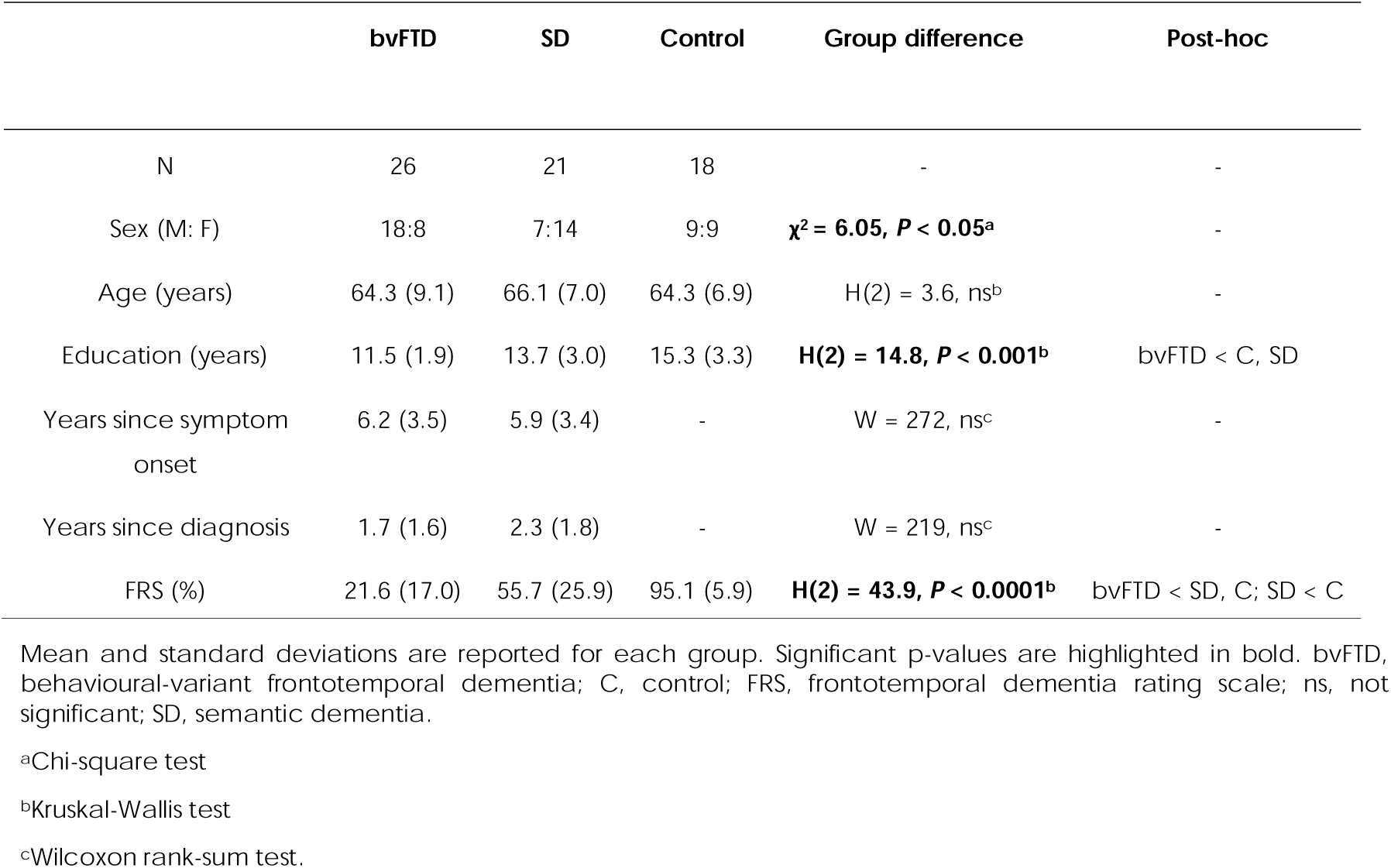
Demographic and disease information.

### Informant questionnaires

#### Cambridge Behavioural Inventory-Revised

The Cambridge Behavioural Inventory-Revised (CBI-R) measures behavioural change in neurodegenerative disorders^12,43^ and includes 45 items which cover ten domains: Memory and Orientation, Everyday Skills, Self-Care, Abnormal Behaviour, Mood, Beliefs, Eating Habits, Sleep, Stereotypic and Motor Behaviours, Motivation. For each item, the informant rates the frequency of the behaviour/functional impairment over the past month on a five-point Likert scale or responds N/A if not applicable. A percentage score for each domain is calculated, where higher scores indicate increased frequency of behavioural change.

#### Apathy-Motivation Index-Caregiver version

The Apathy-Motivation Index-Caregiver version (AMI-CG) is an informant questionnaire designed to measure apathy in neurological patients.^44^ There are 18 items covering three apathy subtypes: Behavioural Activation, Social Motivation and Emotional Sensitivity. For each item, the informant rates how appropriately the behaviour describes the thoughts and behaviours of the patient from five response options ranging from ‘completely true’ to ‘completely untrue’. A score is derived for each apathy domain by averaging item scores, where higher scores indicate greater levels of apathy. The caregiver version was used, as cognitively impaired participants may lack the necessary insight to provide a reliable self-report (e.g., Klar et al.^44^). All controls and a subset of the FTD cohort (bvFTD = 21, SD = 15) also completed the original self-report version of the AMI^45^ to allow a direct comparison of self versus informant ratings.

#### Cambridge Questionnaire for Apathy and Impulsivity Traits

The Cambridge Questionnaire for Apathy and Impulsivity Traits (CamQUAIT) is a 15-item informant questionnaire designed to measure apathy and impulsivity, developed and validated specifically in the context of syndromes associated with frontotemporal lobar degeneration.^46^ Informants rate the frequency of behaviours over recent weeks from four response options. Scores are calculated for two domains – “Motivation and Support” (CamQUAIT-M) and “Impulsivity and Challenging Behaviours” (CamQUAIT-C) where higher scores indicate increased frequency of behaviour change.

### Neuropsychology

Participants completed a battery of neuropsychological tests.^39^ Tasks assessed comprehension of multiple types of social concepts, including famous people,^47,48^ abstract social concepts,^25,34,49–52^ emotions,^53,54^ social norms understanding and sarcasm detection.^55^ Where possible, ‘non-social’ comparator tasks were included, matched for lexical frequency, specificity and imageability.^56^ General semantic memory was assessed using the modified picture version of the Camel and Cactus test (CCT) and naming tasks from the Cambridge Semantic Memory Test Battery,^57–60^ a synonym judgement task^59,61^ and the 30-item Boston Naming Test.^62,63^ Global cognition was assessed using the Addenbrooke’s Cognitive Examination-Revised (ACE-R), a dementia screening tool with five subscales: Attention and Orientation, Memory, Language, Fluency and Visuospatial Function^64^. The Brixton Spatial Anticipation Test^65^ and Raven’s Coloured Progressive Matrices Set B^66^ were included as tests of executive function.

### Structural MRI

A subset of the FTD cohort (bvFTD = 15, SD = 18) and 35 age-matched healthy controls had a T1-weighted 3T structural MRI scan on a Siemens PRISMA, University of Cambridge using an MPRAGE sequence. Of these, 29 participants (bvFTD = 1, SD = 12, control = 16) were scanned at the MRC Cognition and Brain Sciences Unit (TR = 2000ms, TE = 2.85ms, TI = 850ms), and 39 (bvFTD = 14, SD = 6, control = 19) were scanned at the Wolfson Brain Imaging Centre (TR = 2000ms, TE = 2.93ms, TI = 850ms). Raw data were converted to the Brain Imaging Dataset format^67^ and preprocessed using the Computational Anatomy Toolbox version 12 in SPM 12.^68^ Images were segmented into grey matter, white matter and cerebrospinal fluid, modulated, and normalised to MNI space using geodesic shooting.^69^ Normalised grey matter images were spatially smoothed using a 10mm FWHM Gaussian kernel.

Voxel-based morphometry (VBM) was conducted to explore grey matter volume differences between groups. Separate general linear models were fitted for each contrast, with age, ICV and scanner site as covariates, and independent t-tests conducted to compare groups. An explicit mask was used, based on a method recommended for VBM of severely atrophic brains.^70^ Significant clusters were extracted using a cluster-level threshold of *P*(FDR) < 0.05, based on an initial voxel-level threshold of *P* < 0.001. Results were visualised using the xjView toolbox (https://www.alivelearn.net/xjview) and brain regions labelled using the Automated Anatomical Labelling Atlas 3.^71^

### Statistical analysis

Behavioural data were analysed using the ‘rstatix’ package^72^ in R studio version 4.0.3^73^ and IBM SPSS version 28. Normality and equality of variance were assessed by Shapiro-Wilk tests and Levene’s test, respectively. If data were normally distributed then one-way ANOVAs and post-hoc Tukey’s range tests were conducted if there was equality of variance across groups, whereas Welch ANOVAs and post-hoc Games Howell tests were conducted where variances were not equal. Where data were not normally distributed, Kruskal-Wallis tests with post-hoc Dunn’s tests were conducted. A *P* < 0.05 threshold was used to determine statistical significance.

### Frequency of behavioural changes in frontotemporal dementia

Informant-based Likert scale ratings in neurodegenerative disorders may be influenced by symptom duration. For example, caregivers may overestimate the frequency/magnitude of behavioural features initially when these may be noticeably florid and distressing relative to a premorbid baseline. Conversely, caregivers may acclimatise to behaviours over time and thus begin to underestimate their frequency and/or magnitude. Moreover, the emergence of apathy or motor deficits may mitigate the expression of other challenging behaviours. To account for the temporal evolution of the clinical syndromes, we explored the prevalence of individual behaviours in FTD regardless of their frequency. First, for each CBI-R domain, a patient was binarised as ‘impaired’ if they had a rating above ‘never’ or ‘not impaired’ otherwise. Second, each patient was binarised as ‘impaired’ (i.e. item frequency rated more than ‘never’) or ‘not impaired’ (i.e. item frequency rated ‘never’) on each CBI-R item (*n* = 45). Differences in the prevalence of behaviours in bvFTD versus SD were explored using χ^2^ tests.

### Extracting the magnitude and dimensions of behaviour change in FTD

CamQUAIT and AMI data were missing for one bvFTD participant, meaning 1.4% of the total FTD data were missing. For the FTD cohort only, raw scores for each questionnaire domain were z-scored and missing data were imputed using probabilistic principal component analysis.^74,75^ This method requires the number of extracted principal components to be pre-specified and so k-fold cross-validation was conducted to determine the optimum number of components, based on the solution with the lowest root mean squared error for held out cases over 1000 permutations.^76^ Two principal component analyses (PCA) were used in the analyses. In the first, PCA with varimax-rotation was conducted on the questionnaire domains to extract the underlying behavioural dimensions of variation in FTD. An initial PCA extracted four principal components which explained 78.3% of the total variance (see Supplementary Fig. 2 for the factor loadings). One bvFTD participant had an extreme outlier factor score (4.05) on the third principal component, which was reflected as interpreting *psychosis.* This participant was the only case in the sample to display prominent psychotic features, and this component disappeared when this participant was removed from the PCA (the other three components remained stable). Consequently, this participant was excluded and the final PCA was re-run with *n* = 46. The results from this final PCA are reported below and used in all further analyses. Sampling adequacy and suitability of the data for the PCA was assessed using the Keiser-Meyer-Olkin (KMO) test and Bartlett’s test of sphericity. The number of principal components to extract was determined using the elbow method on the scree plot of eigenvalues.^77^ Factor scores for each participant were calculated using the regression method and groups were compared using independent t-tests.

### The neuropsychological and neuroanatomical correlates of behaviour change in FTD

A second, separate varimax-rotated standard PCA was conducted on the FTD neuropsychological data. This PCA extracted three components, labelled for ease of reference and interpretation as: (1) *FTD severity*, (2) *semantic memory* and (3) *executive function* (Supplementary Fig. 3).^39^ Note that such labelling is approximate and implies differential weighting rather than exclusivity of features across components. The SD group had significantly lower factor scores (i.e. poorer performance) on the *semantic memory* component compared with bvFTD (*t* = 5.38, *P* < 0.0001), whereas the bvFTD group had significantly lower scores than SD on the *executive function* component (*t* = 3.97, *P* = 0.0002) (Supplementary Fig. 4).

The association between neuropsychological performance and behaviour change was explored by fitting separate forced-entry linear multiple regression models with factor scores on each component extracted from the informant questionnaire PCA as the dependent variable, and the three neuropsychological components (*FTD severity, semantic memory, executive function)* as predictors. In the FTD cohort only (*n* = 32), voxel-based correlations^78^ were conducted to determine the regions of grey matter intensity associated with factor scores on the informant questionnaire PCA-derived behavioural dimensions. A linear regression model was fitted with each factor score as the dependent variable, and age, ICV and scanner site included as covariates. There were no significant clusters for any correlations using an initial voxel-level threshold of *P* < 0.001. Therefore, we applied a slightly more lenient voxel-threshold of *P* < 0.005, whilst keeping a cluster-level threshold of *P*(FDR) < 0.05.

### BvFTD versus SD discrimination

Logistic regression was conducted to determine the ability of each informant questionnaire PCA-derived behavioural dimension to discriminate between bvFTD and SD. This was compared with the discriminative ability of the three neuropsychological components (described above). Discriminative ability (as a diagnostic forced two-choice classification) was expressed by receiver operator characteristic (ROC) curves with “area under the curve” as the summary metric.

## Results

### Grey matter volume differences between groups

The VBM results align closely with the expected distribution of frontotemporal atrophy in FTD and in each syndrome (Fig. 1 and Supplementary Table 1). The bvFTD group had reduced grey matter volume, which was maximal in the prefrontal cortex, and extended to the temporal lobes, insula, parietal lobe and cerebellum. The SD group had reduced grey matter volume in the bilateral ATLs, maximal at the temporal pole, with additional loss in the posterior temporal cortex, prefrontal cortex and insula. Comparisons between bvFTD and SD revealed reduced volume in the bilateral ATLs in SD, with no significant clusters emerging in the reverse contrast.

**Figure 1.**
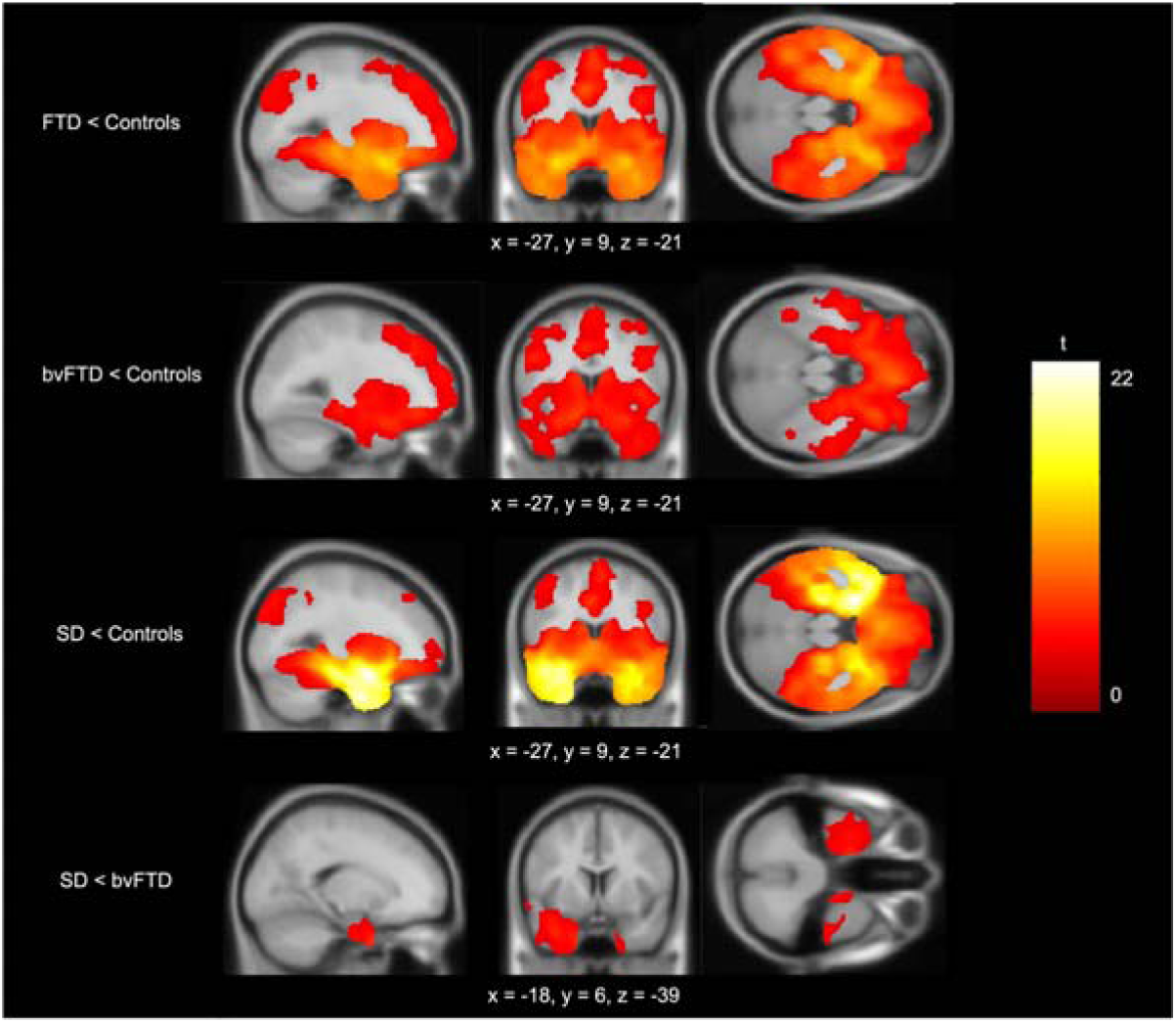
Voxel-based morphometry results. Rows display regions of reduced grey matter volume in each patient group compared to age-matched controls. The bottom row shows regions of reduced grey matter volume in SD compared to bvFTD. Groups were compared using independent t-tests, with age, intracranial volume and scanner site included as covariates. Images are thresholded using a cluster-level threshold of *P*(FDR) *< 0.05* (after an initial voxel-level threshold of *P* < 0.001). Significant clusters are overlaid on the MNI avg152 T1 template. Co-ordinates are reported in Montreal Neurological Institute space.

### Behavioural changes in FTD

Table 2 and Fig. 2A display group average total scores on each questionnaire domain. As expected, the bvFTD group had significantly higher scores (i.e. more severe behaviour change) than controls across every domain. These main effects were not driven solely by the bvFTD sample, however - the SD group also had significantly higher scores than controls on every domain apart from CBI-R Self-Care (*P* = 0.15), CBI-R Beliefs (*P* = 0.09), AMI-CG Emotional Sensitivity (*P* = 0.18) and CamQUAIT-C (*P* = 0.30).

**Figure 2.**
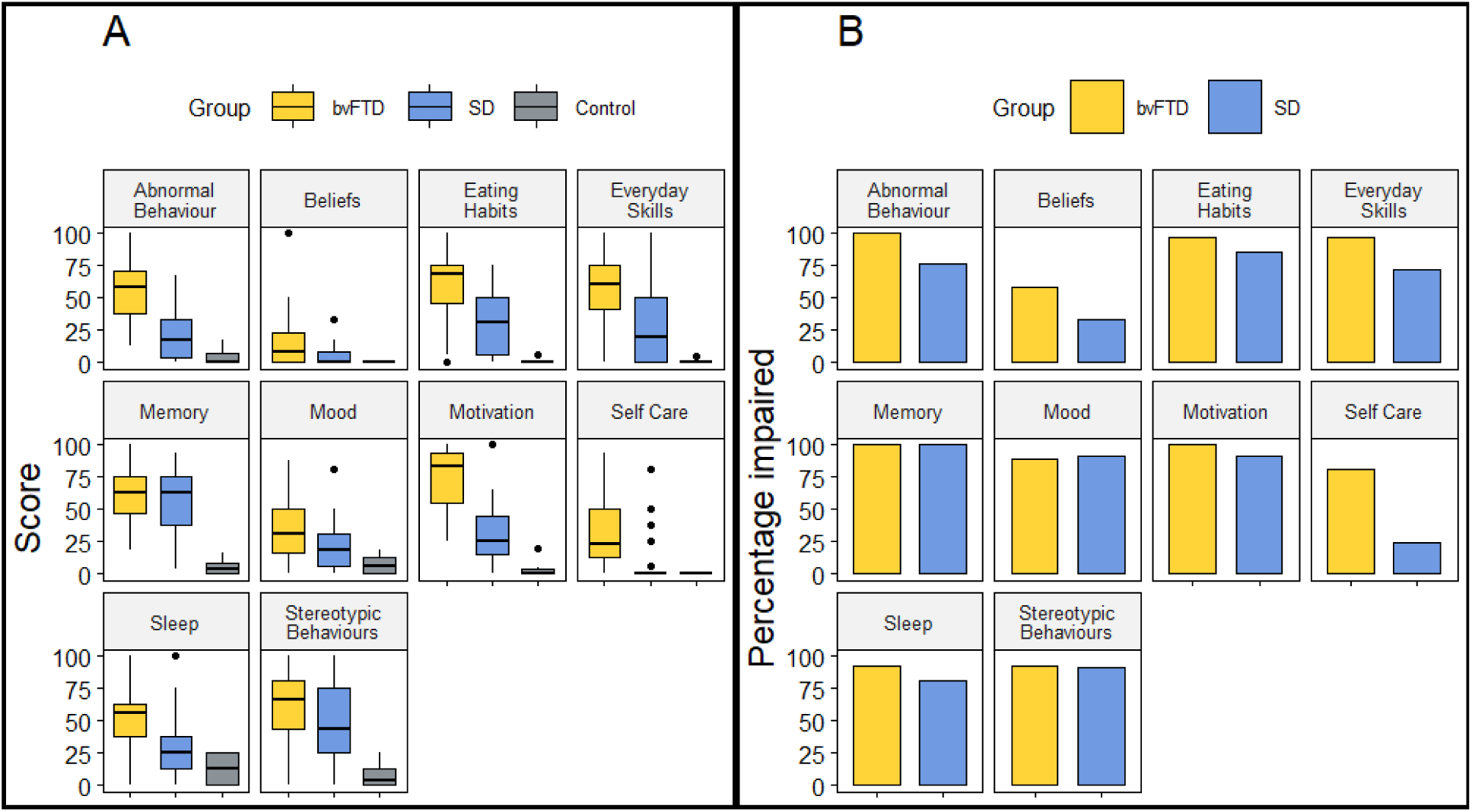
Scores across each CBI-R domain. **(A)** Average total scores across each CBI-R domain in each group. **(B)** The percentage of bvFTD and SD participants impaired on each CBI-R domain.

**Table 2.** Average total scores on each Informant Questionnaire Domain.

| Questionnaire domain | bvFTD | SD | Control | Group effect | Post-hoc |
| --- | --- | --- | --- | --- | --- |
| CBI-R Memory and Orientation (%) | 59.7 (22.9) | 56.1 (24.8) | 5.2 (5.7) | H(2) = 36.7, $P < 0.0001$ | bvFTD, SD > C |
| CBI-R Everyday Skills (%) | 58.7 (27.7) | 29.0 (31.8) | 0.3 (1.2) | H(2) = 36.3, $P < 0.0001$ | bvFTD, SD > C |
| CBI-R Self-Care (%) | 33.9 (30.5) | 9.5 (21.5) | 0.0 (0.0) | H(2) = 29.2, $P < 0.0001$ | bvFTD > C |
| CBI-R Abnormal Behaviour (%) | 55.4 (25.7) | 21.2 (22.1) | 4.2 (6.2) | H(2) = 36.3, $P < 0.0001$ | bvFTD, SD > C |
| CBI-R Mood (%) | 33.4 (23.4) | 21.7 (19.0) | 6.9 (6.7) | H(2) = 18.4, $P < 0.001$ | bvFTD, SD > C |
| CBI-R Beliefs (%) | 14.5 (22.4) | 4.4 (8.2) | 0.0 (0.0) | H(2) = 16.0, $P < 0.001$ | bvFTD > C |
| CBI-R Eating Habits (%) | 60.6 (27.4) | 31.3 (24.0) | 0.7 (2.0) | H(2) = 39.3, $P < 0.0001$ | bvFTD, SD > C |
| CBI-R Sleep (%) | 50.0 (25.7) | 30.4 (28.9) | 11.8 (10.9) | H(2) = 19.8, $P < 0.0001$ | bvFTD, SD > C |
| CBI-R Stereotypic and Motor Behaviours (%) | 60.3 (30.3) | 47.3 (34.7) | 6.9 (8.3) | H(2) = 25.5, $P < 0.0001$ | bvFTD, SD > C |
| CBI-R Motivation (%) | 71.5 (25.6) | 32.1 (25.7) | 2.2 (4.9) | H(2) = 44.0, $P < 0.0001$ | bvFTD, SD > C |
| AMI-CG Behavioural Activation (4) | 3.0 (0.9) | 1.5 (1.0) | 0.5 (0.5) | H(2) = 37.0, $P < 0.0001$ | bvFTD, SD > C |
| AMI-CG Social Motivation (4) | 2.6 (0.8) | 2.1 (0.8) | 1.1 (0.8) | F(2,61) = 21.0, $P < 0.0001$ | bvFTD, SD > C |
| AMI-CG Emotional Sensitivity (4) | 3.0 (0.9) | 1.6 (1.0) | 1.1 (0.6) | H(2) = 29.0, $P < 0.0001$ | bvFTD > C |
| CamQUAIT-M (27) | 20.5 (5.7) | 12.8 (6.0) | 4.7 (3.1) | H(2) = 38.8, $P < 0.0001$ | bvFTD, SD > C |
| CamQUAIT-C (18) | 8.4 (3.9) | 4.2 (3.3) | 2.7 (1.9) | F(2,61) = 17.9, $P < 0.0001$ | bvFTD > C |

Mean and standard deviations are reported for each group. AMI-CG, Apathy-Motivation Index-Caregiver version; bvFTD, behavioural-variant frontotemporal dementia; CamQUAIT, Cambridge Questionnaire for Apathy and Impulsivity Traits; CBI-R, Cambridge Behavioural Inventory Revised; C, control; SD, semantic dementia.

Post-hoc pairwise comparisons between FTD subtypes revealed that the bvFTD group had significantly higher scores on the following CBI-R domains: Everyday Skills (*P* = 0.005), Self-Care (*P* = 0.0003), Abnormal Behaviours (*P* = 0.0006), Eating Habits (*P* = 0.007), Sleep (*P* = 0.04) and Motivation (*P* = 0.002). The bvFTD group also had increased ratings of apathy, with higher scores on the Behavioural Activation (*P* = 0.002) and Emotional Sensitivity (*P* = 0.0002) AMI-CG domains, and both CamQUAIT subscales (CamQUAIT-M, *P* = 0.002; CamQUAIT-C, *P* = 0.0002). There were no differences on the AMI-CG Social Motivation domain (*P* = 0.07) or on the following CBI-R domains: Memory and Orientation (*P* = 0.74), Mood (*P* = 0.12), Beliefs (*P* = 0.09) and Stereotypic and Motor Behaviours (*P* = 0.26). These results demonstrate that people with FTD are impaired across a wide range of behaviours, and this is not selective to bvFTD but is true in SD too (although milder on average).

In each CBI-R domain, the percentage of ‘impaired’ bvFTD patients was above 50% (Fig. 2B). This was also true in SD, except for Self-Care (23.8%) and Beliefs (33.3%). There was a significantly higher proportion of bvFTD patients impaired on the Everyday Skills (χ^2^ = 5.61, *P* = 0.02), Self-Care (χ^2^ = 15.25, *P* < 0.0001) and Abnormal Behaviours (χ^2^ = 6.93, *P* = 0.008) domains. There were no significant differences on the remaining CBI-R domains (Supplementary Table 2).

The percentage of participants impaired on each individual CBI-R item is reported in Supplementary Table 3. For each item (*n* = 45), χ^2^ tests were conducted to explore whether particular behaviours were more prevalent in one FTD subtype than the other. Twenty-five out of 45 (55.6%) χ^2^ tests were significant, and in every single situation, this was due to a significantly higher proportion of impaired bvFTD participants. There were no instances where the opposite was true, i.e. impaired behaviours significantly more frequent in SD. In other words, we detected no “behavioural double dissociations”.

### Association between self- and caregiver-ratings of apathy

Despite large differences in AMI-CG ratings between FTD and controls, there were no differences between groups on the self-rated version of the AMI for Behavioural Activation (*H*(2) = 2.1, *P* = 0.36), Social Motivation (*F*(2,51) = 0.76, *P* = 0.48) or Emotional Sensitivity (*F*(2,51) = 0.03, *P* = 0.97). The correlation between AMI-CG and AMI scores for each group is displayed in Fig. 3. Self- and informant-ratings of apathy were positively correlated in controls (Behavioural Activation; *r* = 0.71, *P* = 0.001, Social Motivation; *r* = 0.58, *P* = 0.01, Emotional Sensitivity; *r* = 0.5, *P* = 0.04). In contrast, there was less concordance between self- and informant ratings in the two FTD subgroups. There were no significant associations in bvFTD (Behavioural Activation; *r* = 0.17, *P* = 0.47, Social Motivation; *r* = 0.25, *P* = 0.28, Emotional Sensitivity; *r* = 0.23, *P* = 0.32). There were significant correlations between SD patients and informant ratings for Social Motivation (*r* = 0.53, *P* = 0.04) and Emotional Sensitivity (r = 0.6, P = 0.02) but not for Behavioural Activation (*r* = 0.23, *P* = 0.41). These findings highlight the discrepancy between the perspective of the patient and the caregiver in FTD, particularly in bvFTD^44^ (and thus why it is important to collect informant reports in the clinic; see Discussion).

**Figure 3.**
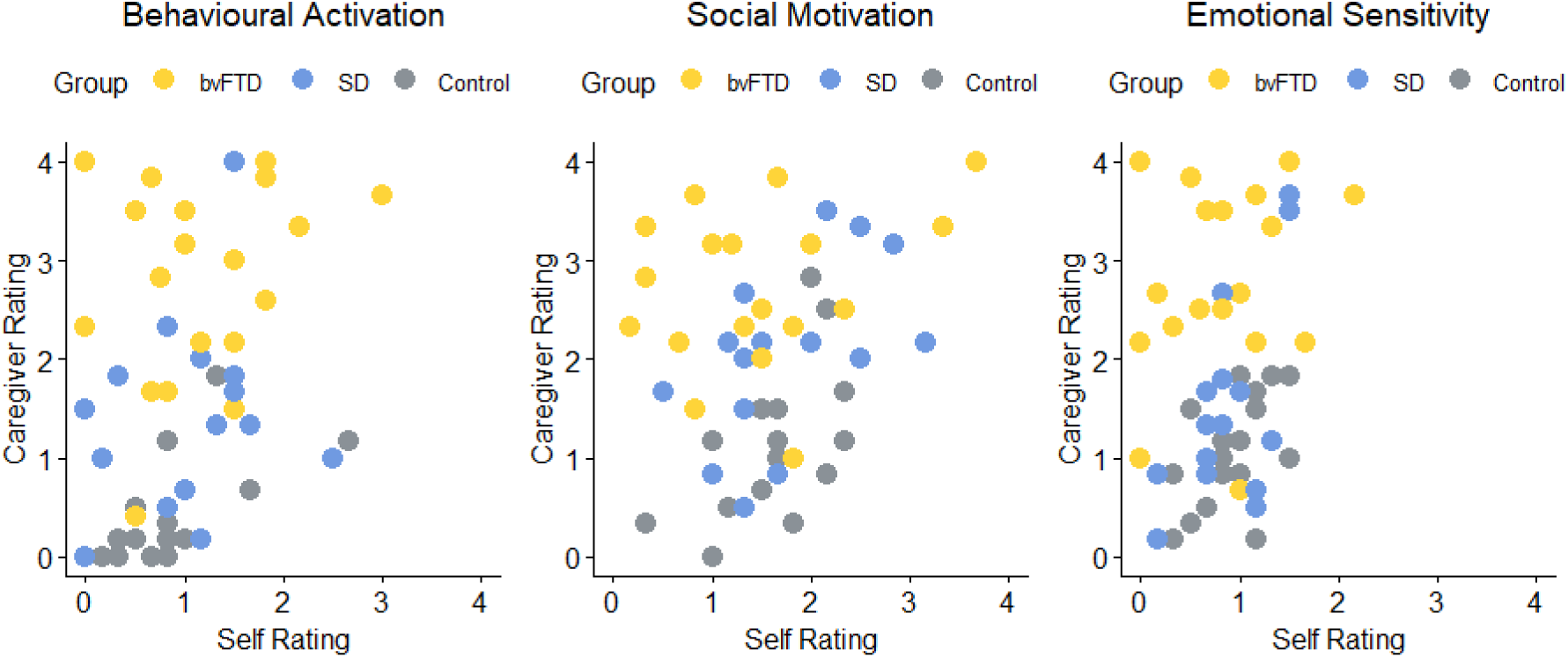
Association between self and informant ratings of apathy. Data points represent scores on the self-rated AMI plotted against scores on the AMI-CG for each AMI domain.

### Extracting the dimensions of behavioural change in FTD

The PCA conducted on the informant questionnaire data had a KMO statistic of 0.75, indicating meritorious sampling adequacy^79^ and Bartlett’s test for sphericity was significant (*P* < 0.0001) indicating presence of at least some common factors in the covariance matrix. Visual inspection of the scree plot indicated three principal components, which explained 77.7% of the total variance. Factor loadings for each questionnaire domain and factor scores for each participant are displayed in Fig. 4. For the full details of the neuropsychology PCA, see Rouse et al.^39^

**Figure 4.**
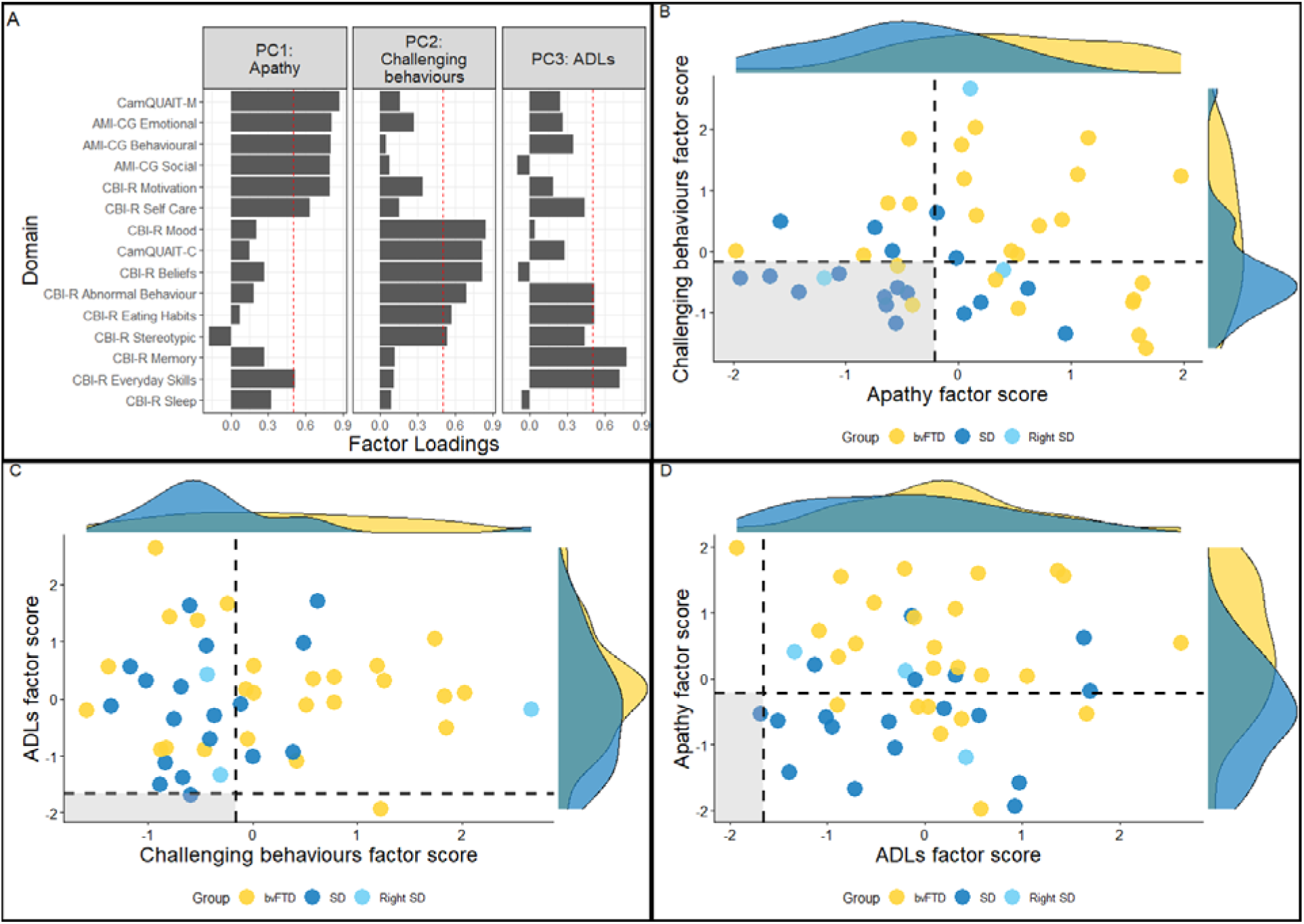
PCA loadings and factor scores. **(A)** Factor loadings for each informant questionnaire domain. Dashed vertical lines indicate the factor loading cut-offs (>0.5). **(B)** PC1 (*apathy*) plotted against PC2 (*challenging behaviours*). **(C)** PC2 (*challenging behaviours*) plotted against PC3 (*ADLs*). **(D)** PC3 (*ADLs*) plotted against PC1 (*apathy*). The dashed lines indicate the factor score of a hypothetical participant scoring 1.96 standard deviations below the control average on each task, and the shaded regions show the regions of preserved performance.

The first principal component had an eigenvalue of 6.96 and explained 46.42% of the total variance. The Motivation, Self-Care and Everyday Skills CBI-R domains, CamQUAIT-M and the three AMI-CG domains loaded positively on this component. Accordingly, this component was labelled *apathy.* The bvFTD group had significantly higher factor scores than SD on this component (*t* = 3.70, *P* = 0.0006). The second principal component had an eigenvalue of 2.38 and explained 15.8% of the total variance. The Mood, Beliefs, Abnormal Behaviours, Eating Habits and Stereotypic and Motor Behaviours CBI-R domains, and the CamQUAIT-C loaded positively on this component. This component was labelled *challenging behaviours.* Again, bvFTD patients had significantly higher factor scores than SD on this component (*W* = 354, *P* = 0.04). The third principal component had an eigenvalue of 1.27 and explained 8.44% of the total variance. The Abnormal Behaviours, Eating Habits, Memory and Orientation and Everyday Skills CBI-R domains loaded positively on this component, and thus the component was labelled *Activities of Daily Living (ADLs).* There were no differences in factor scores between bvFTD and SD on this component (*t* = 1.20, *P* = 0.24).

### BvFTD versus SD discrimination

Receiver operating characteristic curves showing the bvFTD versus SD discriminative ability of each behavioural and neuropsychological component are displayed in Fig. 5. *Semantic memory* had the highest predictive accuracy (AUC = 84.1%) followed by *executive function* (AUC = 78.8%) and *apathy* (AUC = 78.5%). There was poor discriminative accuracy from *challenging behaviours* (AUC = 67.4%), *FTD severity* (AUC = 55.5%) and *ADLs* (AUC = 61.1%). When combined, *semantic memory* and *executive function* had excellent predictive accuracy (AUC = 95.1%) while *apathy, challenging behaviours* and *ADLs* combined had good predictive accuracy (AUC = 83.1%).

**Figure 5.**
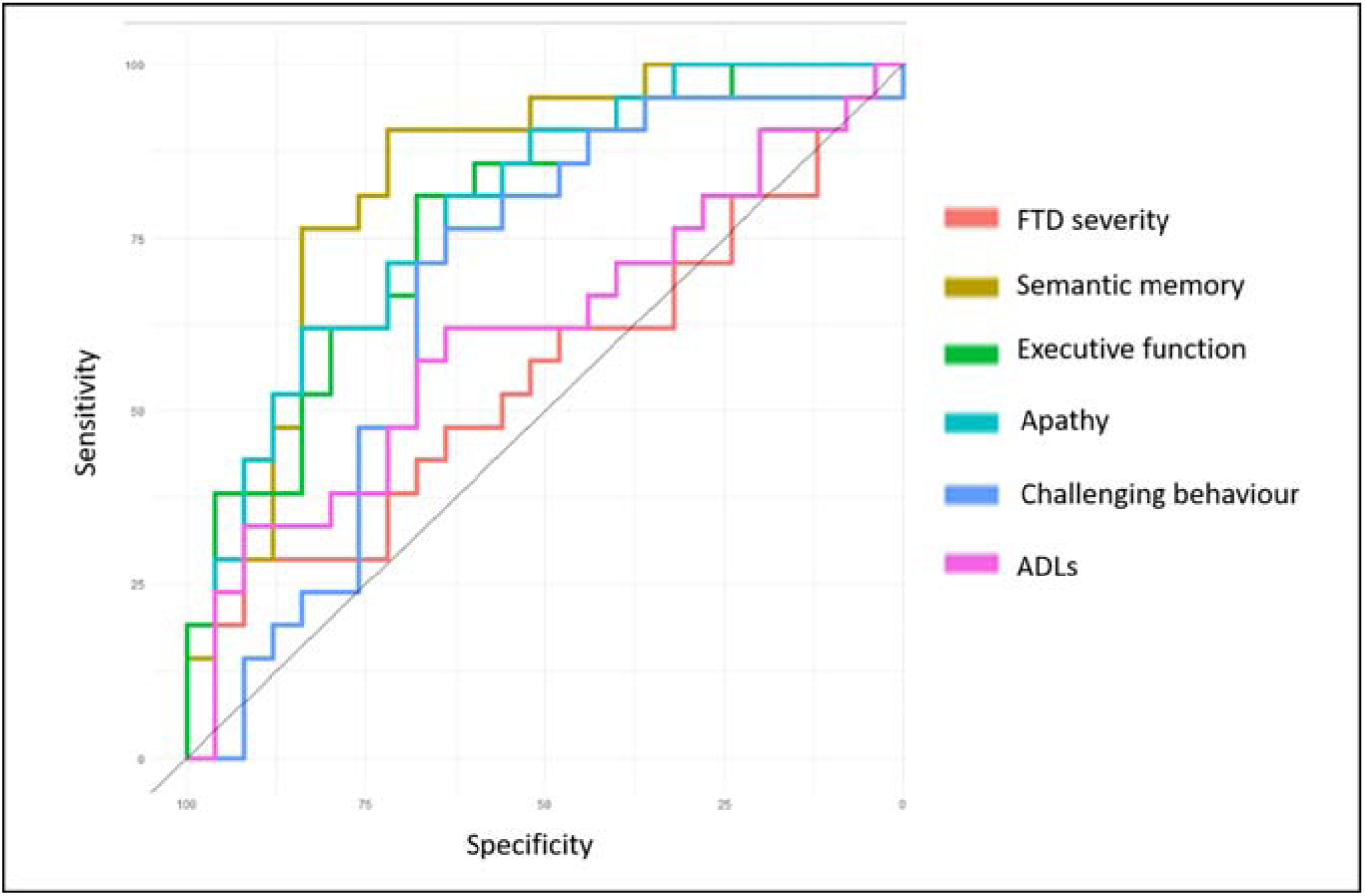
Receiver Operating Characteristic curves distinguishing between bvFTD and SD using the neuropsychological and behavioural components.

### The neuropsychological and neuroanatomical correlates of behaviour change in FTD

A linear multiple regression model with the three neuropsychology components as predictors (*FTD severity*, *semantic memory*, *executive function*) was significant for *apathy* factor scores (*F*(3, 42) = 5.25, *P* = 0.004) with *executive function* the only significant individual predictor (*t* = −3.51, *P* = 0.001, standardized beta = −0.46). The negative beta value indicates that higher levels of apathy were associated with poorer status of executive function. To investigate which specific aspects of executive function were most related to apathy, partial correlations were calculated between each of the three tasks that loaded on the *executive function* factor and *apathy* factor scores, whilst controlling for the other two tasks. *Apathy* factor scores were significantly correlated with performance on the Brixton Spatial Anticipation Test (*r* = −0.56, *P* = 0.001) but not with the Ravens (*r* = −0.03, *P* = 0.90) or TASIT (*r* = 0.06, *P* = 0.76). The model was significant for *ADLs* factor scores (*F*(3, 42) = 5.04, *P* = 0.0001), where *FTD severity* was only significant individual predictor (*t* = −3.18, *P* = 0.003, standardized beta = - 0.42). The negative beta value indicates that increased impairments in ADLs were associated with increased levels of FTD severity. The model was not significant for the *challenging behaviours* component (*F*(3, 42) = 1.85, *P* = 0.15).

Voxel-based correlational analysis detected no regions of grey matter that were associated with factor scores on the *apathy* or *challenging behaviours* component. However, when a measure of global atrophy was added as a covariate, then significant clusters emerged for *apathy* in the dorsal anterior cingulate cortex (Brodmann area 24), supplementary motor area and precuneus. Higher factor scores on the *ADLs* component were negatively associated with grey matter volume in the medial prefrontal cortex, precentral gyri and left insula (Fig. 6 and Supplementary Table 4). A similar set of brain regions was associated with total atrophy and indeed, when total atrophy was included as a covariate in the analysis, then no regions remained for *ADLs*.

**Figure 6.**
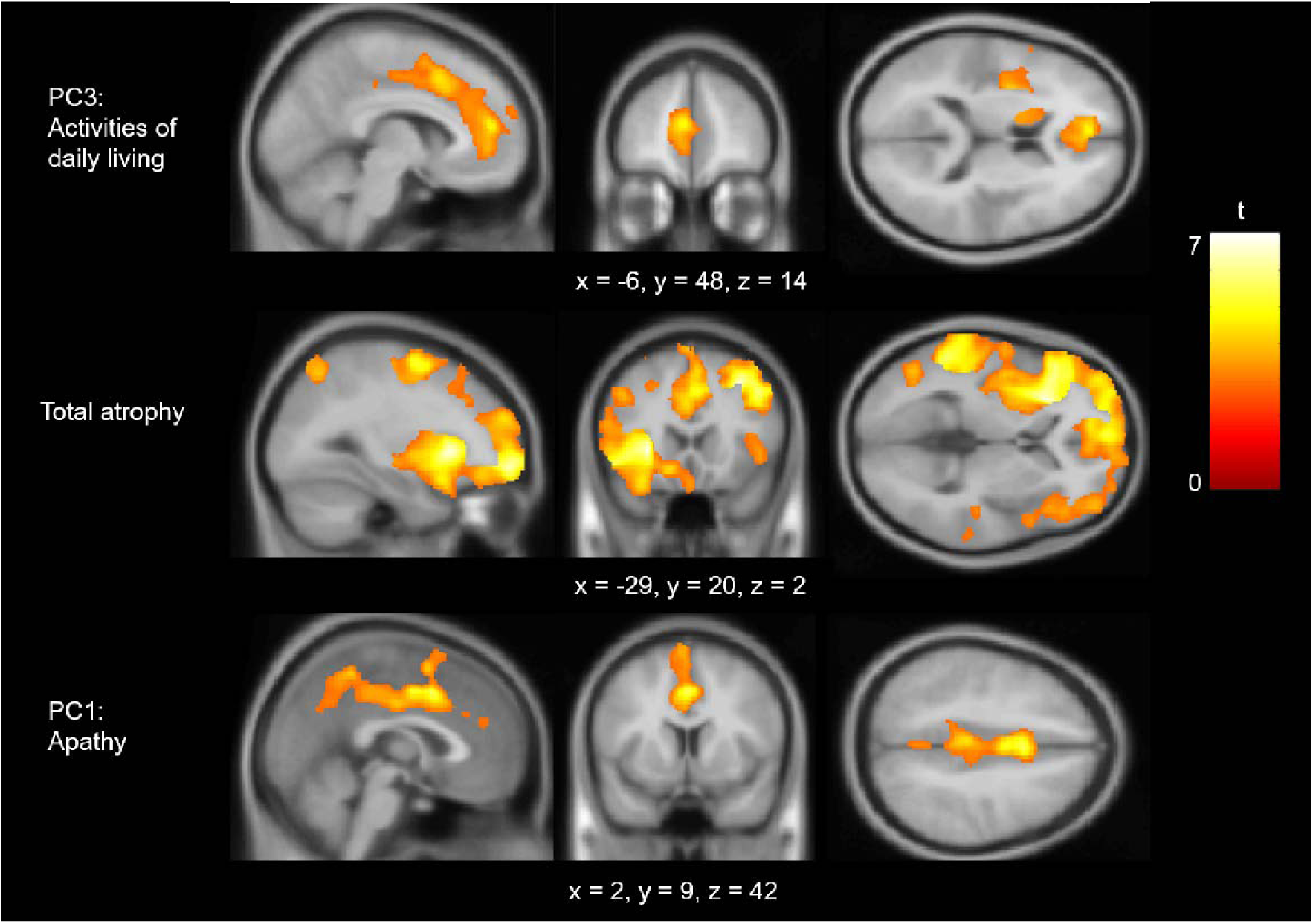
Regions of grey matter volume associated with factor scores. Multiple linear regression models were fitted with each factor as the main effect and age, intracranial volume and scanner site included as covariates. Images are thresholded using a cluster-level threshold of *P*(FDR) < 0.05 (after an initial voxel-level threshold of P < 0.05). Significant clusters are overlaid on the MNI avg152 T1 template. Co-ordinates are reported in Montreal Neurological Institute Space.

## Discussion

Behavioural changes are a common manifestation of frontotemporal dementia; they are a defining feature of bvFTD and are also common in SD.^9,10,12,80^ This study addressed two clinical conundrums, each with important implications. First, are there clear qualitative distinctions between the behavioural profiles in bvFTD and SD? We confirmed the frequency and dimensionality of abnormal behaviours in FTD, with *quantitative* rather than *qualitative* differences between bvFTD and SD. For discrimination of bvFTD versus SD, neuropsychological measures of semantic memory and executive function were much more powerful than behavioural change. We also found that there was a large discrepancy between patients’ self-ratings of apathy *versus* informant ratings, highlighting the importance of the caregiver’s perspective when measuring behavioural change in FTD and for effective evaluation in diagnostic clinics.^81^

Second, what are the roles of the prefrontal cortex and anterior temporal lobe in supporting social behaviour? The transdiagnostic approach, including intermediate cases, reveals the underlying dimensions of behavioural change, and it is the individual expression of these dimensions that was used to study neuroanatomical correlates of FTD behaviour and neuropsychology rather than a traditional binary group comparison. Apathy was a major dimension in FTD, and apathy severity was associated with impaired executive function and anterior cingulate cortex atrophy in both bvFTD and SD. No association was found between behavioural changes and levels of semantic knowledge or ATL grey matter volume. In the following sections, we discuss these findings and their implications, including in relation to the emerging concept of a ‘right temporal variant FTD’– currently a highly debated topic in the field.^82^

### Do behavioural profiles in bvFTD and SD differ quantitatively or qualitatively?

People with bvFTD displayed the expected wide range of behavioural and social disturbances.^5,83^ Behavioural change was common in SD too. Indeed, across CBI-R domains, a high percentage of FTD patients displayed a degree of impairment, in contrast to age-matched controls who were at floor-level. Taken together, these results highlight the sensitivity of informant questionnaires for detecting behaviour changes in FTD and reinforce that both bvFTD and SD patients have abnormal scores across every behavioural domain. The behavioural overlap mirrored the radiological overlap - there was a degree of bilateral ATL volume loss in bvFTD and a degree of prefrontal volume loss in SD, in line with previous neuroimaging comparisons.^41,84^ This confirms the absence of an absolute neuroanatomical division between bvFTD and SD. Instead, each patient has correlated atrophy in multiple regions and occupies a different point in a frontotemporal multidimensional atrophy space. Patients (beyond their initial presentation) often display intermediate phenotypes and express diagnostic criteria for more than one syndrome.^85^ This clinical overlap reflects systematic, graded variations across FTD rather than absolute, mutually exclusive categories. Accordingly, the use only of categorical comparisons seems to be suboptimal for disentangling the precise functions of the ATL and prefrontal regions. Therefore, we also adopted a transdiagnostic approach to FTD and applied multivariate analytics sensitive to the heterogeneity in FTD to model the graded behavioural and cognitive variations and then identify their neuroanatomical underpinnings.

Group-level diagnostic-based comparisons revealed higher levels of apathy, abnormal behaviour, changed eating habits, impaired everyday skills and self-care in bvFTD compared to SD. In contrast, there were no differences in stereotypic behaviours, mood or abnormal beliefs. Although there were many individual behaviours more common in bvFTD, there were no specific behaviours more common in SD. This finding contrasts with some previous studies that have identified distinct behavioural profiles in bvFTD and SD.^9,13^ For example, Snowden et al. reported that, although obsessive behaviours were common in both FTD subtypes, there was a more ‘compulsive’ quality to these behaviours in SD (e.g., clock watching). Here we found that, although compulsive behaviours were indeed highly prevalent in SD, they were even more frequent in bvFTD. Taken together, the lack of any behavioural double dissociations coupled with the broad group-level differences suggests that the behavioural signatures of bvFTD and SD (at least those captured by the questionnaires used) do not differ *qualitatively,* but rather *quantitatively* and *unidirectionally* (bvFTD>SD).

An important clinical implication of contrastive behavioural signatures in bvFTD and SD is improved diagnostic classification. This is particularly salient for disease modifying treatments, as bvFTD and SD are associated with different underlying neuropathologies (TDP-43 type C in SD^86^ and heterogeneous pathology in bvFTD).^87^ Consequently, accurate diagnosis is vital for clinical trial stratification, to ensure that any participant in a trial actually has the proteinopathy the drug is targeting. We found that, although apathy was a good discriminator between bvFTD and SD, behavioural measures had poorer discriminative ability than two neuropsychological measures; *semantic memory* (SD<bvFTD) and *executive function* (bvFTD<SD) and was most powerful when the two neuropsychological components were combined (AUC = 0.95). Previous studies have shown that bvFTD and SD can be clearly distinguished using neuropsychology, with a double dissociation between semantic memory (impaired in SD) and executive function (impaired in bvFTD).^88,89^ This finding implies that, rather than chasing subtle behavioural differences (which appear to be primarily quantitative rather than qualitative in nature), neuropsychological assessments of semantic memory and executive function should be considered the discriminative ‘gold standard’ at least in terms of bedside testing or when neuroimaging is not available. According to current consensus criteria, a possible bvFTD diagnosis requires a neuropsychological profile of executive deficits with relative sparing of episodic memory and visuospatial function.^5^ Based on our findings, we suggest that relatively preserved semantic memory should be included as an important neuropsychological criterion for bvFTD.

### The roles of the prefrontal cortex and anterior temporal lobes in supporting behaviour

The informant questionnaire PCA extracted three behavioural components: *apathy*, *challenging behaviours* and *ADLs*. Apathy is a core feature of FTD; it is a diagnostic criterion for bvFTD^5^ and is also common in SD.^90^ Apathy is considered a multidimensional construct and distinct subtypes have been proposed, each associated with different neural circuitry.^91,92^ In the current study, behavioural, social and emotional apathy domains were highly intercorrelated and co-loaded onto the same component, indicating that all three domains are concurrently affected by FTD.^93^ Direct comparisons between FTD subtypes revealed increased severity of apathy in bvFTD, in keeping with this feature as a core diagnostic criterion.^5^ There was evidence for increased apathy in SD too – indeed the majority of patients (61.9%) had apathy factor scores outside of the control-defined normality cut-off (see Fig. 4). These findings highlight the prevalence of apathy in FTD and its occurrence across the FTD spectrum. Apathy can be difficult to distinguish from depression as they both include features such as loss of interest and anhedonia.^94^ However, in our study the mood domain of the CBI-R did not co-load with the apathy measures but loaded onto a statistically orthogonal component (*challenging behaviours).* This suggests that the motivational changes found in this study were not due to affective changes, in keeping with other studies which have found apathy and depression to be dissociable in FTD.^95^

The CS-SC model proposes that the impaired social behaviour in FTD can result from damage to two distinct yet interactive components: (i) *social-semantic knowledge*, underpinned by the bilateral ATLs, and (ii) *social control*, including selection, evaluation, decision-making and inhibition supported by prefrontal cortical regions. A core hypothesis from the model is that atrophy in medial prefrontal regions will cause deficits in the ability to control and regulate social-semantic knowledge effectively, to guide appropriate and adaptive social behaviours.^8^ By taking a transdiagnostic approach, we were able to reveal the underlying behavioural dimensions across the FTD clinical spectrum and show that anterior cingulate cortex atrophy is associated with increased levels of apathy, aligning with the predictions of the CS-SC framework.

Increased apathy was associated with poor executive function, a finding which replicates previous FTD studies,^96,97^ and which can emerge years before conversion from presymptomatic to symptomatic states in genetic FTD.^98^ When the executively-loading tasks were analysed separately, performance on the Brixton Spatial Anticipation Test was the only task that was significantly correlated with apathy, potentially indicating a more specific relationship between apathy and certain aspects of executive function (e.g., the ability to adapt flexibly to rule changes and inhibit previous response strategies). It was not possible from our study to determine the causal relationship between apathy and executive function, however a previous study reported that apathy predicts executive cognitive decline in presymptomatic genetic bvFTD.^98^

Voxel-based correlational analysis revealed that apathy severity in FTD was negatively correlated with grey matter volume in the anterior cingulate cortex. This was true not only of the bvFTD sample (classically associated with anterior cingulate cortex atrophy) but in SD too, indicating that (a) the increased levels of apathy in bvFTD>SD reflects the predominance of prefrontal atrophy in the former condition and (b) the apathy in SD is a consequence of pathology spreading into medial prefrontal areas (rather than a distinct neurocognitive process associated with ATL atrophy, for example). Atrophy or hypometabolism in the anterior cingulate cortex has been strongly linked with apathy in FTD^99–102^ as well as in other neurodegenerative disorders such as Alzheimer’s disease^103,104^ and Parkinson’s disease.^105^ Moreover, anterior cingulate cortex lesions cause severe apathy and abulia.^106^ The neurocognitive mechanism of social control deficits underlying apathy in FTD can be explained by a predictive coding framework as a ‘failure of active inference’ due to reduced precision of prior expectations, leading to failures in correctly adapting actions to the environment and thus diminished goal-directed behaviour.^8,107^ In support of this hypothesis, apathy is associated with reduced prior precision in both healthy participants and people with Parkinson’s disease.^108,109^ The anterior cingulate cortex may be the anatomical substrate of goal priors, or potentially underpin a hub for the integration of prior expectations with sensory inputs. Apathy is a multidimensional construct, where even theorised subdomains such as ‘emotional sensitivity’ or ‘social motivation’ might encompass multiple behavioural subcomponents. It is possible that two people with FTD might exhibit ‘apathy’ for different mechanistic reasons, which would raise the question of whether the syndromes should be regarded as equivalent on this behavioural dimension. Although apathy had a common neuroanatomical correlate in bvFTD and SD, future studies that utilise functional neuroimaging^110^ and ancillary physiologically informed techniques^111,112^ may be able to deconstruct the complex behavioural changes that are called apathy and disinhibition.

A key hypothesis of the CS-SC framework is that the impaired behaviour in SD is predominantly due to a degradation of social-semantic knowledge following bilateral ATL atrophy.^8^ Here we found no association between impaired social-semantic knowledge and behavioural change in FTD. How does this fit with the predictions of the CS-SC framework? First, our social-semantic battery already contains tasks which provide direct measures of social abilities (e.g., emotion recognition, person recognition, sarcasm detection) and the SD patients were impaired on these and more so than the bvFTD subgroup.^39^ Thus these direct assessments do detect social changes, and we have formally shown that they are very highly correlated with both general (non-social) semantic impairments and atrophy in the ATL bilaterally.^39^ Second, unlike some of these direct measures, it seems possible that questionnaires such as the CBI-R may miss these more “semantically driven” aspects of behavioural change, and instead are more sensitive to deficits in prefrontal-based “social control” processes such as apathy or disinhibition. If correct, then the development of better-targeted informant questions, sensitive to these aspects of behaviour change, including formal assessment of behavioural change associated with SD in earlier studies^9,10,113^ is an important avenue for future research.

### Implications for the ‘right temporal variant of FTD’

FTD patients with R>L ATL atrophy often present with behavioural changes; this clinical observation is routinely observed in specialist clinics^114,115^ (although there are exceptions).^116^ In recent years, efforts have been made to characterise and define the right ATL temporal variant, motivated in part because of the high clinicopathological correlation with TDP43-opathy rather than tauopathy, and in part because the existing criteria for svPPA do not include the associated behaviour changes.^14^ This has led to several proposals of diagnostic criteria and an appropriate label for these patients, including the ‘right temporal variant of FTD’,^35^ and ‘semantic behavioural-variant FTD’.^33^ An international working group has been formed, with the aim to define a cohesive clinical phenotype for this syndrome, driven by the lack of uniform consensus criteria and nomenclature.^117^ A multi-centre retrospective analysis of 360 FTD patients with predominant right ATL atrophy found that the most common symptoms at initial presentation were: compulsive behaviours, disinhibition/socially inappropriate behaviour, naming/word-finding difficulties, memory deficits, apathy, loss of empathy, prosopagnosia, and problems recognising and altered reactions to taste, bodily sensations, smell and sound.^117^ However, despite R>L asymmetry in all cases, only four cases had selective right ATL atrophy. This complicates the localisation of function of these features to the right ATL. For example, many of the behavioural features listed above (reduced empathy, apathy, compulsive behaviours, social disinhibition) are also common in bvFTD,^5^ and even the behaviours considered to be associated more with ‘right temporal variant FTD’ than with bvFTD (e.g., rigid preoccupations and narrowed food preferences) can be seen in L>R SD patients too.

One mechanism behind the behavioural changes in R>L SD is a degradation of social-semantic knowledge following right ATL atrophy.^33,52,117^ However, a recent study found that there were no differences in social-semantic knowledge between L>R and R>L SD patients, and that both general semantic knowledge and social-semantic knowledge were associated with *bilateral* ATL volume.^39^ L>R and R>L patients had overlapping neuropsychological profiles, without highly selective social-semantic deficits in R>L ATL cases. A similar pattern was found in the current study - we did not find any evidence for behavioural disturbances specific to those with R>L ATL atrophy. Rather, R>L and L>R patients were highly overlapping in terms of their position along the behavioural dimensions and exhibited bilateral levels of ATL atrophy (although we note that coverage of every possible relevant behavioural feature was not possible). In summary, the data from FTD suggest that social-semantic knowledge is part of a broader conceptual system underpinned by the bilateral ATL. This is consistent with three lines of evidence from other patient groups and healthy participants. First, selective right ATL resection for temporal lobe epilepsy does not cause a selective impairment for social concepts or lead to behavioural changes.^39,118^ Second, distortion-corrected or distortion-reducing fMRI studies in healthy participants have found bilateral ventrolateral ATL activation for both social and non-social concepts.^49,51^ Finally, transcranial magnetic stimulation to the left *or* right ATL generates a transient disruption to social-semantic decision making.^50^

Why then, do people with R>L ATL atrophy consistently present with behavioural problems? Group studies have found that R>L SD patients typically have more temporal lobe atrophy overall than L>R,^31,41^ and increased prefrontal atrophy.^119^ Based on our CS-SC model, there are at least two (non-mutually exclusive) alternative explanations for the increased behavioural change in R>L SD. First, R>L SD patients have greater total ATL volume loss bilaterally, leading to a greater degradation of social-semantic knowledge which is important for appropriate social behaviour.^8^ Second, the increased behavioural changes in R>L SD result from their correlated atrophy in prefrontal areas important for social control, such as the orbitofrontal cortex and anterior cingulate cortex.

Our findings have implications for the nosological status of the ‘right temporal variant of FTD’. We conceptualise SD as a *unitary-yet-graded* disorder, where SD is an umbrella term for a neurodegenerative disorder encompassing both L>R (svPPA) and R>L (‘right’ SD, ‘right temporal variant FTD’), where L>R and R>L cases represent spectrum points of the same disease. The *unitary-yet-graded* model of SD is supported by several key findings. First, although atrophy may be asymmetric early in SD, hypometabolism tends to be more symmetrical.^120^ Second, L>R and R>L cases become increasingly similar over time and merge into the same clinical syndrome as atrophy rapidly spreads into the contralateral ATL.^36,119,121,122^. Third, L>R and R>L ATL atrophy is associated with TDP-43 type C pathology, suggesting these cases constitute a single disease.^123^ Clinical heuristics and educational material may reasonably highlight the differences between bvFTD and right temporal variant FTD or right SD, not least because of the differences in the risk of genetic mutations, motor neuron disease and parkinsonism. However, we recommend that a research agenda motivated by mechanistic insights and aspirations for disease-modifying treatments does not get distracted by attempts to impose binary diagnostics on L>R versus R>L syndromes, as if they represented distinct diseases.

### Limitations and future directions

People with FTD often lack insight into their changed behaviours^15,19,124–126^ and may have cognitive deficits that cause unreliable response strategies^127^ and violations of the assumptions underlying questionnaires and self-report forms (e.g., of consistent and meaningful responding).^127^ Indeed, this was highlighted by the discrepancy between patient and caregiver reports of apathy in the current study. Informant ratings/interviews are therefore a common and important method for assessing behavioural changes in both clinical and research settings. However, there are multiple possible factors to consider when interpreting informant ratings. For example, there is evidence that informant ratings are influenced by caregiver burden.^128^ Additionally, there may be an interaction between informant ratings of behaviour and the time since the behaviour’s onset, such that in the early stages of the disease the obvious change from premorbid baseline ‘magnifies’ any behavioural changes and leads to disproportionately high ratings. Indeed, this possibility motivated the parallel analyses of CBI-R behaviour severity/frequency versus behaviour presence in the current study (see above).

The plethora of behavioural features associated with FTD makes it extremely challenging for a single study to fully capture the spectrum of clinically relevant features. Indeed, it was this challenge that motivated our decision to assemble a battery of informant questionnaires rather than rely on a single standard tool. Nevertheless, we acknowledge that the current study did not cover all the possible behavioural domains that may contribute to bvFTD versus SD phenotypic discrimination, such as anosognosia,^129^ empathy loss,^33^ and aberrant reward behaviours.^27^ Relatedly, the factor loadings and interpretation of a PCA are dependent on the tasks entered, meaning that if a dataset fails to include crucial behavioural features present in the patient population studied, the resultant PCA will not derive a principal component for this clinically relevant behavioural dimension. Our conclusion of quantitative not qualitative differences between FTD syndromes pertains to the behaviours sampled by our measures, but a crucial mission for future research is to incorporate a more comprehensive survey of all relevant behaviours. This includes diverse ethnic and linguistic groups, which may differ from the UK regarding the tolerance and response to social norm violations in people with FTD.^130,131^

Our cross-sectional study design meant that many participants were several years into their dementia; the average time since symptom onset was around six years. As typical of previous FTD cohort studies, there were overlapping radiological profiles between bvFTD and SD with atrophy extending beyond the initial respective atrophy centres (Fig. 1). It is therefore possible that discriminative behavioural signatures between bvFTD and SD exist early on but soon disappear as atrophy spreads and clinical phenotypes become increasingly similar, and thus not detected in the current study^17,85,132^ A critical avenue for future research is to track similarity/differences in behaviours longitudinally to explore how behavioural phenotypes merge or diverge. In addition, studies could retrospectively analyse informant questionnaires from previous clinic visits, to investigate whether selective behavioural features exist in FTD subtypes in the earlier stages of the disease.

The PCA-based approach in this study may have obscured some more specific relationships between social-semantic impairments and particular aspects of behaviour change, such as socially inappropriate behaviour or “disinhibition”. However, without inclusion of more specific tests of social executive functions, it remains difficult to differentiate between prefrontal versus temporal contributions to inappropriate social behaviour. Future studies could relate informant ratings of behavioural changes with performance on social executive tasks used in previous FTD studies, which assess the ability to resolve social dilemmas^133^ and social decision making.^134^

## Data availability

Due to the limits of the ethics approval for these patient studies, the raw data cannot be openly shared. Requests for anonymised data can be addressed to the senior author and may require a data transfer agreement.

## Acknowledgements

We thank all the patients and their caregivers for giving up the time to take part in the study. We also thank Dr. Thomas Cope, Dr. Sian Thompson and Dr. Sofia Toniolo for their help with FTD recruitment. For the purpose of open access, the UKRI-funded authors have applied a CC BY public copyright licence to any Author Accepted Manuscript version arising from this submission.

## Funding

M.A.R is supported by the Medical Research Council (SUAG/096 G116768). J.B.R is supported by the Medical Research Council (MC_UU_00030/14; MR/T033371,1), Wellcome Trust (220258), and the NIHR Cambridge Biomedical Research Centre (NIHR203312). M.A.L.R is supported by a Medical Research Council programme grant (MR/R023883/1) and intramural funding (MC_UU_00005/18). The views expressed are those of the authors and not necessarily those of the NIHR or the Department of Health and Social Care.

## Competing interests

The authors report no competing interests.

## Notes

### Competing Interest Statement

The authors have declared no competing interest.

### Author Declarations

The National Research Ethics Service's East of England Cambridge Central Committee gave ethics approval for this work(IRAS project ID: 252986)

### Summary of Updates

The Discussion section now includes implications for the right temporal variant FTD. Additional tables and figures have been added to the Supplementary Material.

