## Supplementary Material for "Behavioural changes in frontotemporal dementia and their cognitive and neuroanatomical correlates"

### SUPPLEMENTARY TABLES

**Supplementary Table 1 Grey matter volumetric differences between groups**

| Contrast | Hemisphere | Number of voxels | Peak MNI co-ordinate | | | Peak MNI co-ordinate region | t-value |
|---|---|---|---|---|---|---|---|
| | | | *x* | *y* | *z* | | |
| *FTD < Controls* | | | | | | | |
| | Bilateral | 263,318 | -27 | 9 | -21 | Temporal pole | 13.53 |
| | Right | 3,069 | 29 | -71 | -50 | Cerebellum | 4.97 |
| *bvFTD < Controls* | | | | | | | |
| | Bilateral | 147,625 | 2 | 36 | -15 | Gyrus rectus | 8.94 |
| *SD < Controls* | | | | | | | |
| | Bilateral | 228,408 | -27 | 3 | -23 | Temporal pole | 21.82 |
| | Right | 3,335 | 29 | -69 | -48 | Cerebellum | 4.73 |
| *SD < bvFTD* | | | | | | | |
| | Left | 9,659 | -18 | 6 | -39 | Temporal pole | 6.23 |
| | Right | 2,448 | 15 | -6 | -38 | Undefined | 5,51 |

bvFTD, behavioural-variant frontotemporal dementia; FTD, frontotemporal dementia; MNI, Montreal Neurological Institute; SD, semantic dementia.

**Supplementary Table 2 Percentage of patients impaired on each CBI-R domain**

| CBI-R Domain | bvFTD | SD | $X^2$ test |
|---|---|---|---|
| Memory and Orientation (%) | 100 | 100 | - |
| Everyday Skills (%) | 96.2 | 71.4 | **$\chi^2 = 5.61$, $P = 0.02$** |
| Self Care (%) | 80.8 | 23.8 | **$\chi^2 = 15.25$, $P < 0.0001$** |
| Abnormal Behaviour (%) | 100 | 76.2 | **$\chi^2 = 6.93$, $P = 0.008$** |
| Mood (%) | 88.5 | 90.5 | $\chi^2 = 0.05$, $P = 0.82$ |
| Beliefs (%) | 57,7 | 33.3 | $\chi^2 = 2.77$, $P = 0.1$ |
| Eating Habits (%) | 96.2 | 85.7 | $\chi^2 = 1.63$, $P = 0.20$ |
| Sleep (%) | 92.3 | 81.0 | $\chi^2 = 1.35$, $P = 0.25$ |
| Stereotypic and Motor Behaviours (%) | 92.3 | 90.5 | $\chi^2 = 0.05$, $P = 0.82$ |
| Motivation (%) | 100 | 90.5 | $\chi^2 = 2.59$, $P = 0.11$ |

Significant p-values are highlighted in bold. bvFTD, behavioural-variant frontotemporal dementia; CBI-R, Cambridge Behavioural Inventory-Revised; SD, semantic dementia.

**Supplementary Table 3 Percentage of patients impaired on each CBI-R item**

| Domain | Item | bvFTD | SD | $X^2$ test |
|---|---|---|---|---|
| Memory and Orientation (%) | Has poor day-to-day memory | 96.2 | 95.0 | $X^2$ = 0.04, $P$ = 0.85 |
| Memory and Orientation (%) | Asks the same questions over and over again | 88.5 | 85.7 | $X^2$ = 0.08, $P$ = 0.78 |
| Memory and Orientation (%) | Loses or misplaces things | 92.3 | 75.0 | $X^2$ = 2.63, $P$ = 0.11 |
| Memory and Orientation (%) | Forgets the names of familiar people | 96.2 | 95.2 | $X^2$ = 0.02, $P$ = 0.88 |
| Memory and Orientation (%) | Forgets the names of objects and things | 92.0 | 100 | $X^2$ = 1.76, $P$ = 0.19 |
| Memory and Orientation (%) | Shows poor concentration when reading or watching television | 88.5 | 81.0 | $X^2$ = 0.52, $P$ = 0.47 |
| Memory and Orientation (%) | Forgets what day it is | 88.5 | 57.1 | **$X^2$ = 5.99, $P$ = 0.01** |
| Memory and Orientation (%) | Becomes confused or muddled in unusual surroundings | 92.3 | 76.2 | $X^2$ = 2.38, $P$ = 0.12 |
| Everyday Skills (%) | Has difficulties using electrical appliances | 88.0 | 38.1 | **$X^2$ = 12.52, $P$ = 0.0004** |
| Everyday Skills (%) | Has difficulties writing letters | 92.0 | 47.6 | **$X^2$ = 11.09, $P$ = 0.0009** |
| Everyday Skills (%) | Has difficulties using the telephone | 92.0 | 42.9 | **$X^2$ = 13.02, $P$ = 0.0003** |
| Everyday Skills (%) | Has difficulties making a hot drink | 48.0 | 9.5 | **$X^2$ = 7.98, $P$ = 0.005** |
| Everyday Skills (%) | Has problems handling money or paying bills | 95.8 | 52.4 | **$X^2$ = 11.45, $P$ = 0.0007** |
| Self Care (%) | Has difficulties grooming self | 73.1 | 23.8 | **$X^2$ = 11.28, $P$ = 0.0008** |
| Self Care (%) | Has difficulties dressing self | 73.1 | 4.8 | **$X^2$ = 22.18, $P$ < 0.0001** |
| Self Care (%) | Has problems feeding self without assistance | 26.9 | 4.8 | **$X^2$ = 4.04, $P$ = 0.04** |
| Self Care (%) | Has problems bathing or showering self | 64.0 | 19.0 | **$X^2$ = 9.39, $P$ = 0.002** |
| Abnormal Behaviour (%) | Finds humour or laughs at things other do not find funny | 92.3 | 42.9 | **$X^2$ = 13.58, $P$ = 0.0002** |
| Abnormal Behaviour (%) | Has temper outbursts | 76.9 | 42.9 | **$X^2$ = 5.71, $P$ = 0.02** |
| Abnormal Behaviour (%) | Is uncooperative when asked to do something | 96.2 | 47.6 | **$X^2$ = 14.39, $P$ = 0.0001** |
| Abnormal Behaviour (%) | Shows socially embarrassing behaviour | 92.0 | 57.1 | **$X^2$ = 7.62, $P$ = 0.006** |
| Abnormal Behaviour (%) | Makes tactless or suggestive remarks | 80.8 | 38.1 | **$X^2$ = 8.95, $P$ = 0.003** |
| Abnormal Behaviour (%) | Acts impulsively without thinking | 96.2 | 52.4 | **$X^2$ = 12.42, $P$ = 0.0004** |
| Mood (%) | Cries | 30.8 | 33.3 | $X^2$ = 0.04, $P$ = 0.85 |
| Mood (%) | Mood | 66.7 | 81.0 | $X^2$ = 1.17, $P$ = 0.28 |
| Mood (%) | Is very restless or agitated | 84.0 | 66.7 | $X^2$ = 1.89, $P$ = 0.17 |
| Mood (%) | Is very irritable | 76.0 | 47.6 | **$X^2$ = 3.95, $P$ = 0.047** |
| Beliefs (%) | Sees things that are not really there | 25.0 | 4.8 | $X^2$ = 3.27, $P$ = 0.07 |
| Beliefs (%) | Hears voices that are not really there | 23.1 | 0.0 | **$X^2$ = 5.56, $P$ = 0.02** |
| Beliefs (%) | Has odd or bizarre ideas that cannot be true | 46.2 | 33.3 | $X^2$ = 0.79, $P$ = 0.37 |
| Eating Habits (%) | Prefers sweet foods more than before | 88.5 | 76.2 | $X^2$ = 1.24, $P$ = 0.27 |
| Eating Habits (%) | Wants to eat the same foods repeatedly | 80.8 | 60.0 | $X^2$ = 2.41, $P$ = 0.12 |
| Eating Habits (%) | Her/his appetite is greater, s/he eats more than before | 73.1 | 23.8 | **$X^2$ = 11.28, $P$ = 0.0008** |
| Eating Habits (%) | Table manners are declining | 84.6 | 38.1 | **$X^2$ = 10.89, $P$ = 0.001** |
| Sleep (%) | Sleep is disturbed at night | 88.5 | 66.7 | $X^2$ = 3.30, $P$ = 0.07 |
| Sleep (%) | Sleeps more by day than before | 69.2 | 42.9 | $X^2$ = 3.31, $P$ = 0.07 |
| Stereotypic and Motor Behaviours (%) | Is rigid and fixed in her/his ideas and opinions | 84.6 | 61.9 | $X^2$ = 3.15, $P$ = 0.08 |
| Stereotypic and Motor Behaviours (%) | Develops routines from which s/he can not easily be discouraged | 76.9 | 66.7 | $X^2$ = 0.61, $P$ = 0.44 |
| Stereotypic and Motor Behaviours (%) | Clock watches or appears pre-occupied with time | 73.1 | 57.1 | $X^2$ = 1.31, $P$ = 0.25 |
| Stereotypic and Motor Behaviours (%) | Repeatedly uses the same expression or catch phrase | 80.8 | 81.0 | $X^2$ = 0.0003, $P$ = 0.99 |
| Motivation (%) | Shows less enthusiasm for his or her usual interests | 88.5 | 52.4 | **$X^2$ = 7.56, $P$ = 0.006** |
| Motivation (%) | Shows little interest in doing new things | 92.3 | 66.7 | **$X^2$ = 4.93, $P$ = 0.03** |
| Motivation (%) | Fails to maintain motivation to keep in contact with friends or family | 92.3 | 66.7 | **$X^2$ = 4.93, $P$ = 0.03** |
| Motivation (%) | Appears indifferent to the worries and concerns of family members | 92.3 | 61.9 | **$X^2$ = 6.41, $P$ = 0.01** |
| Motivation (%) | Shows reduced affection | 96.2 | 57.1 | **$X^2$ = 10.56, $P$ = 0.001** |

Significant p-values are highlighted in bold. bvFTD, behavioural-variant frontotemporal dementia; SD, semantic dementia

**Supplementary Table 4 Regions of grey matter intensity correlating with factor scores**

| | Hemisphere | Number of voxels | Peak MNI co-ordinate | | | Peak MNI co-ordinate region | *t*-value |
|---|---|---|---|---|---|---|---|
| | | | *x* | *y* | *z* | | |
| *PC3 – ADLs* | | | | | | | |
| | Bilateral | 11,025 | -6 | 48 | 14 | Anterior cingulate cortex | 4.72 |
| | Left | 8,871 | -29 | 14 | -2 | Undefined | 4.52 |
| *Total Atrophy* | | | | | | | |
| | Bilateral | 102,982 | -29 | 20 | 2 | Insula | 6.60 |
| | Right | 3,792 | 60 | -42 | -23 | Inferior temporal gyrus | 4.58 |
| *PC1 – Apathy* | | | | | | | |
| | Bilateral | 6,180 | 2 | 9 | 42 | Middle cingulate gyrus | 5.52 |

ADLs, activities of daily living; MNI, Montreal Neurological Institute.

### SUPPLEMENTARY FIGURES

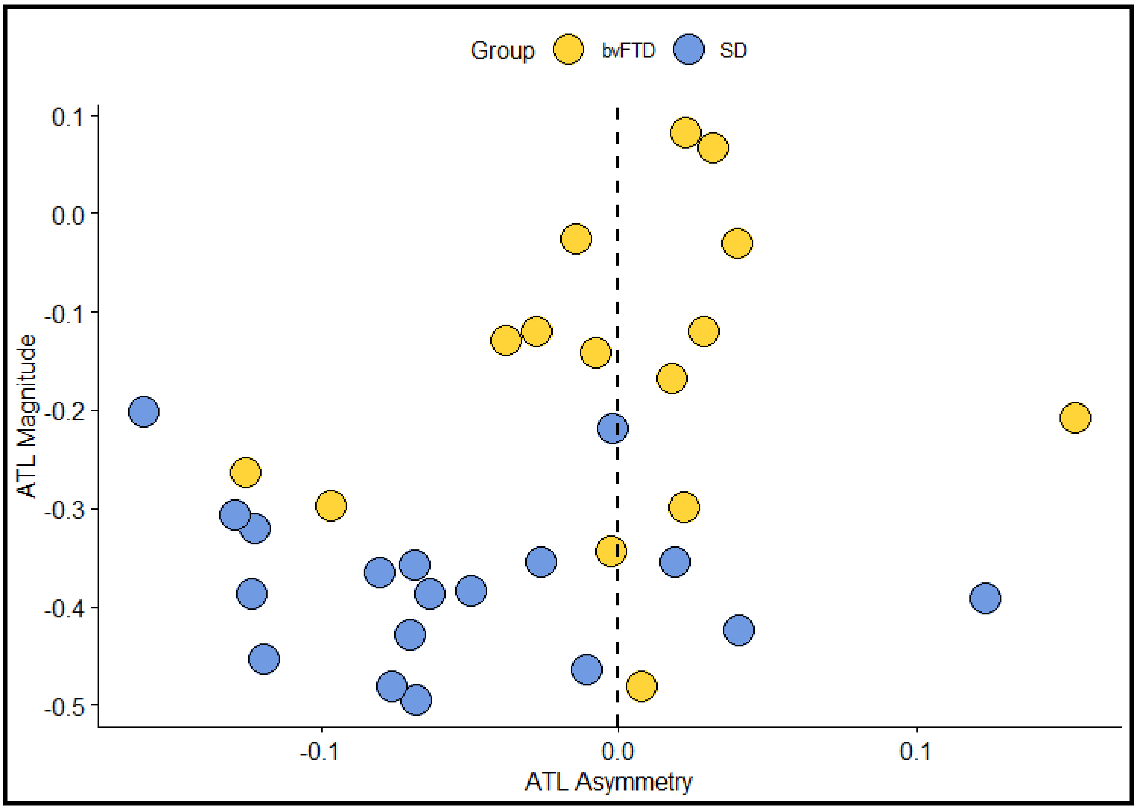


**Supplementary Figure 1. ATL indices for each patient**. Lower magnitude values indicate increased grey matter volume loss. Negative asymmetry values indicate left>right grey matter volume loss whereas positive asymmetry values indicate right>left grey matter volume loss.

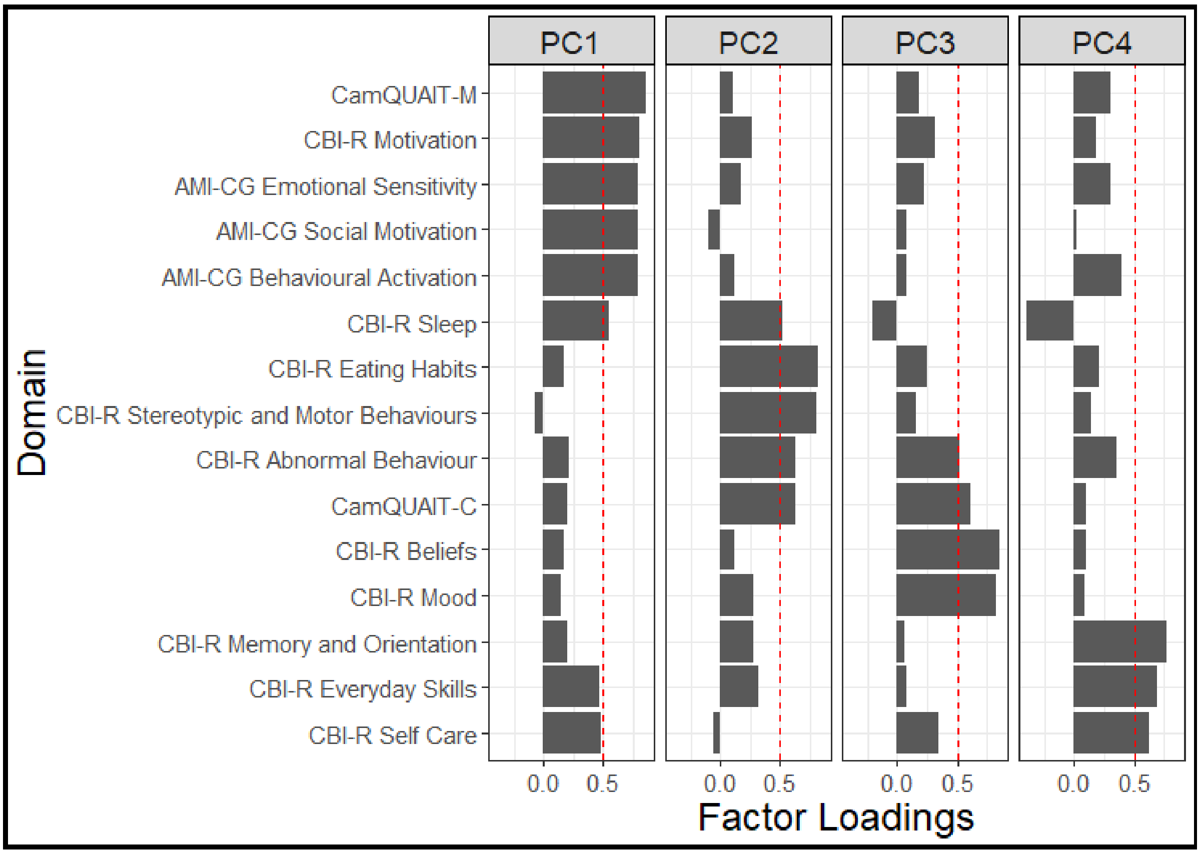


**Supplementary Figure 2**. **Factor loadings of the informant questionnaire domains for the initial PCA**. Dashed vertical lines indicate factor loading cut-offs (>0.5).

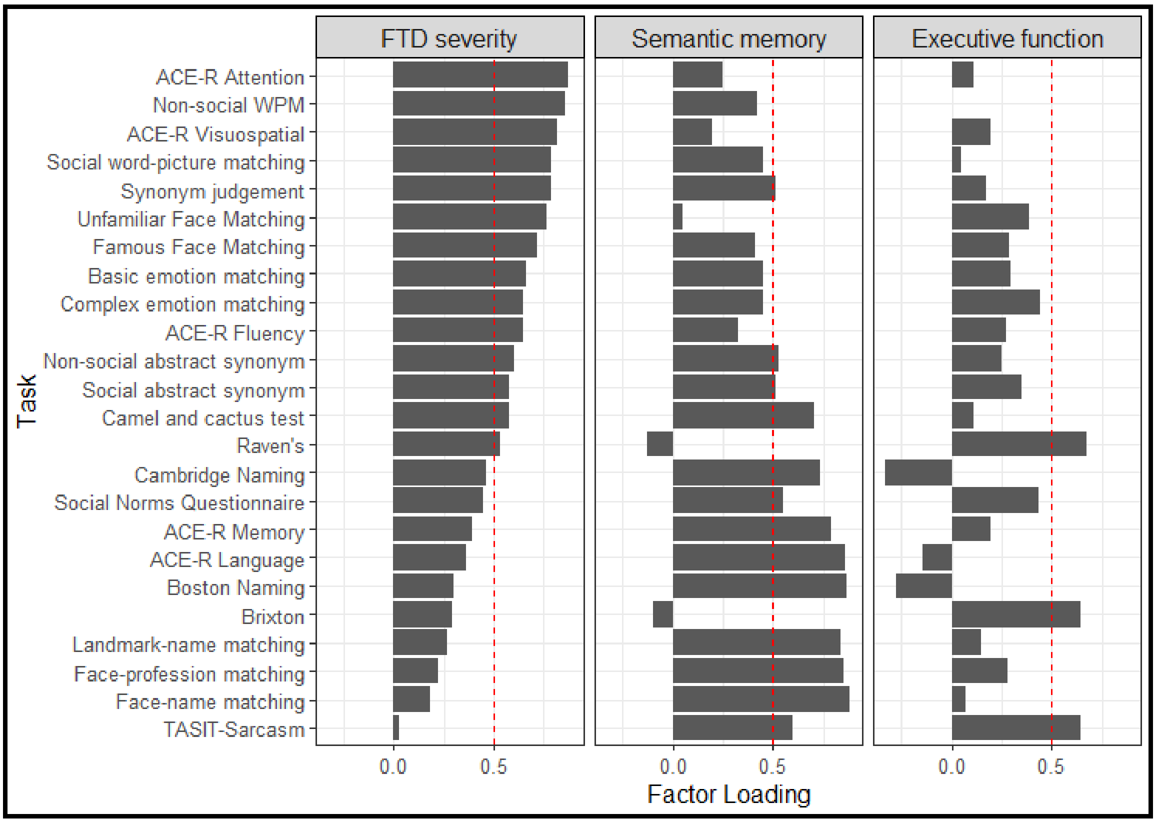


**Supplementary Figure 3**. **Factor loadings of the neuropsychological tasks**. Dashed vertical lines indicate factor loading cut-offs (>0.5).

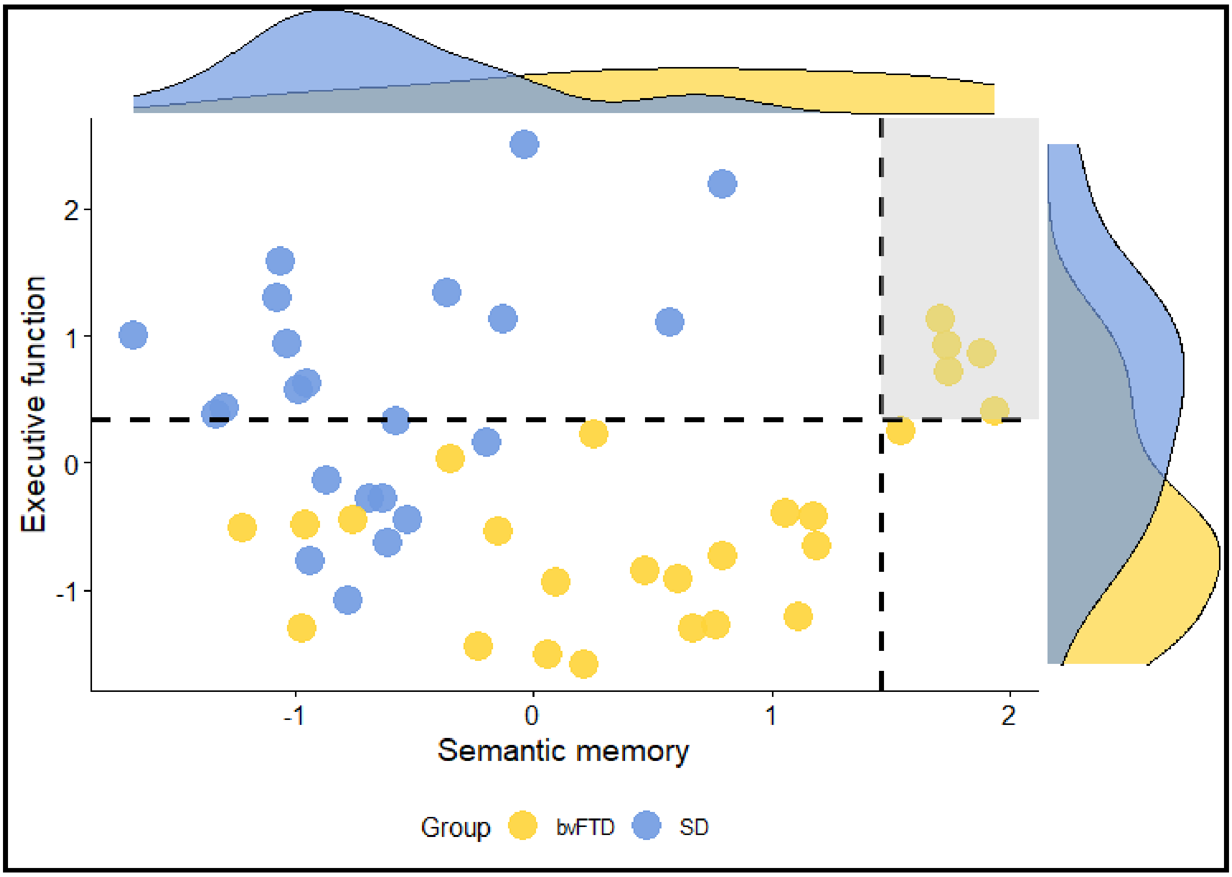


**Supplementary Figure 4. Neuropsychology factor scores.** Dashed lines indicate the factor score of a control scoring 1.96 standard deviations below the control average on each task and the shaded region shows the region of preserved performance.
